# BREATHE: A realist evaluation protocol to understand how smoking cessation services support pregnant women in areas of social deprivation

**DOI:** 10.64898/2026.06.04.26354590

**Authors:** N Carlisle, M Zhang, NAB Simpson, T Stacey

**Affiliations:** Methodologies Division, Florence Nightingale Faculty of Nursing, Midwifery & Palliative Care, King’s College London, London, United Kingdom; Department of Women’s and Children’s Health, School of Medicine, University of Leeds, Leeds, United Kingdom

## Abstract

**Background:** Tobacco smoking during pregnancy increases the risk of preterm birth, small for gestational age (SGA), stillbirth, and longer-term adverse health outcomes. Globally, reducing smoking in pregnancy is a key public health priority, yet the organisation, accessibility, and effectiveness of cessation support varies substantially between countries and healthcare systems. Differences in policy implementation, resource allocation, and integration of cessation services into antenatal care influence uptake and success rates across diverse settings. In England, pregnant women are entitled to free smoking cessation support, however, service delivery varies across regions with mixed efficacy.

While tobacco smoking is more prevalent in deprived communities, there is limited understanding of how, why, for whom, and under what circumstances these services are most effective, particularly in areas of social deprivation, such as the North East and Yorkshire.

**Objective:** To conduct a realist evaluation to understand how smoking cessation services support pregnant women in areas of social deprivation to stop smoking and reduce adverse perinatal outcomes.

**Methods:** This multi-site realist evaluation will be conducted across three NHS maternity services in West Yorkshire, England. The study comprises four iterative stages: (1) development of initial programme theories through realist-informed literature scoping and stakeholder consultation; (2) case study data collection including qualitative interviews with pregnant women (approximately 15-30) and staff (approximately 15-30); (3) analysis of routine anonymised maternity and neonatal electronic data collected over a one-year period; and (4) realist analysis to refine context-mechanism-outcome (CMO) configurations. Qualitative data will be analysed using realist logic supported by NVivo software. Quantitative data will be analysed using descriptive and inferential statistics to explore associations between smoking cessation engagement and perinatal outcomes.

**Ethics and dissemination:** Ethical approval was obtained through the UK Health Research Authority and a Research Ethics Committee prior to study commencement (IRAS 364173; REC reference number 26/SC/0020). Findings will inform recommendations to improve smoking cessation support for pregnant women in deprived areas. Results will be disseminated through peer-reviewed publications, conference presentations, and stakeholder engagement.

## Introduction

### Background

Tobacco smoking during pregnancy remains a significant public health issue, and tobacco use and exposure to second-hand smoke during pregnancy are major and preventable threats to maternal and child health(1). This is recognised globally, with smoking during pregnancy contributing substantially to preventable perinatal morbidity and mortality across high-, middle-, and low-income countries, despite long-standing public health efforts to reduce prevalence (1). Babies born to women who smoke are at increased risk of preterm birth, small for gestational age (SGA), stillbirth, perinatal mortality, and long-term morbidity (2–4). Those living in areas of social deprivation are more likely to smoke at the start of pregnancy and less likely to quit successfully (5,6). These socioeconomic gradients in smoking during pregnancy are consistently observed across many international settings, highlighting the persistent influence of structural inequalities on maternal health behaviours (7).

Across England, 10% of women are known smokers at the start of pregnancy (at their first antenatal appointment, otherwise known as the ‘booking appointment’) (8). By delivery, this rate drops to 4.7% of women (9). However, in the North East of England and Yorkshire, recent data suggest that 5.5% of pregnant women are still smoking at delivery, the highest rate in England (9).

In England, smoking cessation support is offered free of charge to pregnant women (10). However, variation exists in referral pathways, intensity of behavioural support, monitoring practices, and integration within maternity services (11,12). These services constitute complex interventions embedded within broader organisational and social systems (11,13).

While tobacco smoking is more prevalent in deprived communities (ONS, 2023), there is limited understanding of how, why, for whom, and under what circumstances these services are most effective, particularly in areas of social deprivation, such as the North East and Yorkshire (Tatton & Lloyd, 2023; Tatton & Melendez-Torres, 2026). Complex behavioural change interventions, such as smoking cessation interventions, typically involve interacting biological, psychological, and social components and are implemented within dynamic contexts (14). International evidence similarly suggests that the effectiveness of such interventions is highly context-dependent, with outcomes varying according to health system structure, cultural norms, and levels of social inequality (15). Traditional evaluation approaches, such as randomised controlled trials (RCTs), primarily assess whether an intervention is effective. However, this is often insufficient for complex interventions, as the same intervention may produce different outcomes across populations and settings. Therefore, there is a need for research to understand why specific interventions work, do not work, or partially work for different groups of populations (16). Realist evaluation is a theory-driven approach designed to understand how, why, for whom, and under what circumstances interventions generate outcomes.

There are underlying assumptions on how complex service interventions will create outcomes (16,17). In a realist evaluation (which is the study design for BREATHE) the initial step is to extract and develop these assumptions, known as (initial) programme theories. A programme provides a resource, an opportunity, or a constraint - all of which can modify the decision-making process of its anticipated target group (18). It is this decision-making process that establishes which outcomes are achieved. A mechanism refers to the interaction between what a programme offers and how its target audience interprets and responds to it. Understanding and explaining often implicit mechanisms is essential to realist evaluation (16,19). Mechanisms can be triggered in both favourable and unfavourable contexts, leading to intended and unintended outcomes. A programme theory creates a proposed relationship between a context (C), mechanism (M) and outcome (O). This is known as a CMO configuration (16).

Understanding the mechanisms through which smoking cessation services influence behaviour and perinatal outcomes is essential to improving service delivery in socially deprived areas. By examining CMO configurations, this study aims to generate transferable explanations that can inform service improvement across maternity systems.

### Aim

To investigate how smoking cessation services support pregnant women in areas of social deprivation in England to stop smoking.

### Research Question

How, why, for whom, to what extent, and in what contexts do smoking cessation services support pregnant women in areas of social deprivation to stop smoking and reduce adverse perinatal outcomes?

### Objectives

#### Primary objectives

- To identify contexts and mechanisms leading to positive and negative smoking cessation outcomes.
- To understand relationships between contexts, mechanisms, and outcomes.
- To identify intended and unintended consequences of smoking cessation services.
- To develop refined programme theories and recommendations for practice and policy.

#### Secondary clinical objectives

To determine whether smoking cessation services are associated with reductions in:

- Preterm birth (<37 weeks’ gestation)
- Small for gestational age (<10^th^ birthweight centile)
- Stillbirth

## Methods

### Study Design

This study was registered with ISRCTN (ISRCTN18430450) and gained ethical approval (IRAS 364173; REC reference number 26/SC/0020) on 14^th^ January 2026.

This study adopts a realist evaluation design. Whilst realist evaluation is an iterative, non-linear process, this study will follow four research stages (20) :

1. **Stage 1: Development of initial programme theories:** Formulating initial programme theories regarding how to support pregnant women with smoking cessation (using CMO statements for how different component may work) through a realist informed scope of the literature, and discussions with the Project Advisory Team (key stakeholders) to ask for their input to ensure the study is built on realistic and meaningful assumptions.
2. **Stage 2: Case study data collection:** Data will be collected from three case sites, guided by the initial CMO statements, to test the programme theories.
3. **Stage 3: Realist analysis:** Data from the case sites will be analysed using a realist logic of analysis (21) to interrogate the programme theories.
4. **Stage 4: Theory refinement and synthesis:** Synthesising and interpreting the findings will refine the initial theories, leading to an understanding of how, for whom, in which circumstances, and why smoking cessation support works (or does not work). This will inform improvements in smoking cessation services and support the development of recommendations for its use across a range of areas.

The study duration is 22 months (September 2025 to July 2027). Participant recruitment and data collection is anticipated to be completed by December 2026, and results are expected by September 2027. See Figure 1 (Realist Research Cycle adapted from (22).

### Study Setting

The study will be conducted in three NHS maternity services in England:

As they are all situated within West Yorkshire; although these sites serve broadly similar sociodemographic populations, they vary in smoking cessation service configuration.

### Case Study Data Collection: Qualitative interviews

Participants will include:

- Staff involved in smoking cessation pathways (5-10 per site)
- Pregnant women eligible for or engaged with cessation services (5-10 per site)

Interviews will be conducted via telephone or video call.

Women identified as eligible at each site will be approached by the direct care team and provided with a Participant Information Sheet. Demographic, medical, and obstetric history data may be used to select a purposive realist sample to test all relevant programme theories(23). If a woman wishes to participate, informed consent will be obtained by the researcher conducting the interviews. Interviews will take place via telephone or video call at a time convenient to the participant.

Appropriate staff will be contacted to create a purposive realist sample, either via email or approached in person by the direct care team (their colleagues) or the research team. They will be provided with information about the study and offered a Participant Information Sheet. If staff members wish to participate, informed consent will be obtained by the researcher conducting the interviews. Interviews will be conducted via telephone or video call at a time convenient for the participant.

Sampling will follow realist principles, selecting participants based on their relevance to testing specific CMO configurations(20,23).

Interviews will explore experiences of smoking cessation support, perceived facilitators and barriers, contextual influences, and unintended consequences, following a realist interview guide (23).

### Case Study Data Collection: Routine electronic hospital data

Non-identifiable, non-individualised maternity and neonatal data will be collected for a one-year period from each site. The specific data to be analysed will be finalised after the programme theories have been formulated in Stage 1. However, it is expected to include the below (which incorporates the Preterm Birth Core Outcome Set (24)):

- Demographics (age, parity, ethnicity, IMD status, BMI)
- Gestation at booking
- Smoking tobacco status at booking
- Referral to smoking tobacco cessation service
- If attended smoking tobacco cessation service
- Involvement in any smoking cessation initiative
- Smoking status at 36 weeks gestation/time of delivery
- Birth and neonatal outcomes (birth weight, gestational age at birth, sex, admission to neonatal unit)

### Sample Size

Realist evaluation does not require formal power calculations for qualitative sampling. Sampling is driven by relevance and rigour, rather than the quantity of data or sample size alone(20,25). Approximately 30-60 interviews are anticipated across three sites (26), which is comparable to other realist research in maternity care (27,28). Routine hospital data will include all women accessing maternity care during a defined one-year period.

## Data Analysis

### Qualitative analysis

Interviews will be transcribed, anonymised, and analysed using realist logic to test and refine programme theories (20,29). Analysis will focus on identifying CMO configurations. NVivo software will support data management (30).

### Quantitative analysis

Descriptive statistics will summarise smoking prevalence and service uptake. Chi-squared tests, Fisher’s exact tests, and binomial logistic regression will explore associations between smoking cessation engagement and perinatal outcomes (preterm birth and SGA). Quantitative findings will inform refinement of programme theories.

### Ethics Approval and Consent to Participate

Ethical approval was obtained from the UK Research Ethics Committee and the Health Research Authority prior to study commencement (IRAS 364173; REC reference number 26/SC/0020).

Written informed consent will be obtained electronically from all interview participants. Participants may withdraw until 1 October 2026, after which data will have been anonymised and incorporated into theory development.

### Data Management

Data will be processed in accordance with the UK General Data Protection Regulation (GDPR) and Data Protection Act 2018.

Interview recordings will be deleted after transcription and verification. Anonymised transcripts and routine data will be stored securely for 10 years.

### Patient and Public Involvement

A Project Advisory Group including Patient and Public Involvement (PPI) women with lived experience, clinicians, and policy representatives will meet three times a year. The Project Advisory Group will contribute to theory development, interpretation, and dissemination planning.

## Dissemination

Findings will be disseminated through direct feedback to participating sites, peer-reviewed publications, conference presentations, and stakeholder engagement. Participants who request updates will receive a summary of the study findings.

Findings will also be shared with relevant NHS service leads, commissioners, and policymakers to support the development and refinement of smoking cessation support within maternity services. The refined programme theories and recommendations generated through this study may inform future policy and practice relating to smoking cessation support for pregnant women across a range of healthcare settings.

## Authors’ contributions

TS and NS conceived and designed the study, and the original funding proposal. Once funded, NC developed the study protocol with input from TS and MZ. TS, NS, NC and MZ will provide methodological expertise throughout the study. MZ will provide oversight of participant recruitment, data collection and liaison with the study sites. NC wrote the first draft of this manuscript and all authors critically reviewed and approved the final version of the manuscript.

## Data Availability Statement

No datasets are generated at protocol stage. Data generated during the study will be available upon reasonable request, subject to ethical and data protection approvals.

